# A sparse proteomic risk score incorporating plasma MMP12 level improves prediction of abdominal aortic aneurysm

**DOI:** 10.64898/2025.12.16.25341932

**Authors:** Michael G. Clark, Shuai Yuan, Susanna C. Larsson, Michael G. Levin, Jakob Woerner, Dokyoon Kim, Scott M. Damrauer

## Abstract

**Background:** Screening criteria for abdominal aortic aneurysm (AAA) are based on clinical factors, such as age and smoking history, but do not include biological factors that may better reflect disease pathogenesis.

**Objectives:** We sought to determine whether a proteomic risk score (ProRS) incorporating plasma protein abundance could improve prediction of AAA.

**Methods:** We performed a cross-sectional analysis of nearly 37,000 participants in the UK Biobank Pharma Proteomics Project with plasma protein abundance data for 274 cardiometabolic proteins. ProRS models were developed using regularized regression.

**Results:** The generated sparse ProRS contained well-established clinical risk factors as well as a single protein – matrix metalloproteinase 12 (MMP12). Overall performance and discriminatory utility of this model was higher than an identical model without MMP12 (difference in Brier score 2.1 × 10^-4^, 95% CrI 2.0-2.3 × 10^-4^; difference in AUROC 0.021, 95% CrI 0.020 - 0.022). Within the cohort, current AAA screening recommendations applied to 4.6% of the population and captured 30% of cases, whereas screening 4.6% of the population at highest risk by ProRS captured 52% of cases. Among individuals with incident AAA, MMP12 abundance was independently associated with time to rupture or repair (HR 1.86, 95% CI 1.39–2.50, p < 0.001). Additionally, MMP12 level improved discrimination of AAA in an external cohort (SIMPLER; difference in AUROC 0.084, 95% CrI 0.081 - 0.087).

**Conclusions:** A biologically-plausible ProRS incorporating a single matrix metalloproteinase improved prediction of AAA over clinical factors alone. These results may be used to enhance screening strategies for AAA.

## Introduction

Abdominal aortic aneurysm (AAA) is characterized by dilation of the abdominal aorta and can progress to aortic rupture and death if left untreated [1]. Current United States Preventive Services Task Force (USPTF) and Society for Vascular Surgery (SVS) screening recommendations for AAA are based on well-established clinical risk factors – including advanced age, male sex, history of smoking, and family history – and involve a one-time screening ultrasound covered under Medicare by the Screening Abdominal Aortic Aneurysms Very Efficiently (SAAAVE) Act [1, 2]. While current screening recommendations are effective in terms of cost and mortality reduction [3], screening is currently underutilized [4], does not capture all high-risk individuals [5], and has a decreasing yield as the overall prevalence of aneurysmal aortic disease declines [6]. Importantly, current screening criteria do not incorporate biological factors that may more accurately reflect underlying disease pathogenesis than clinical or demographic factors alone.

The availability of large-scale biobanks has enabled discovery of novel genetic associations with AAA, facilitating development of polygenic risk scores [7, 8] and genetics-informed screening strategies [9, 10, 11]. A critique of polygenic risk scoring is that the calculated risk depends only on fixed genetic factors, and therefore does not reflect other important biological and environmental factors that modify disease risk over time. Plasma proteomic profiling has emerged as a complementary approach for risk prediction across a wide range of diseases [12]. Previous work to establish relationships between plasma proteins and AAA has been small in scale [13] or used genetically imputed protein levels as opposed to direct measurement of plasma protein abundance [14, 15]. Nonetheless, these studies have yielded interesting and biologically plausible associations, with a particularly strong signal for proteins implicated in extracellular matrix maintenance and remodeling.

To build upon these results, and to meet the clinical need for improved AAA screening criteria, we leveraged large-scale proteomic data from the UK Biobank to develop and validate a plasma proteomic risk score (ProRS) for AAA. Our objective was to determine whether measurement of plasma protein abundance could be used to enhance AAA risk stratification beyond use of clinical factors alone.

## Methods

### Study Population

The UK Biobank (UKB) is a large-scale population cohort study of approximately half a million adults from across the United Kingdom. Blood plasma protein profiles were generated for a subset of these participants as part of the UK Biobank Pharma Proteomics Project (UKB-PPP) using antibody-based Olink assays [16]. The UKB and UKB-PPP resources received ethical approval from the North West Multi-Centre Research Ethics Committee (reference number 21/NW/0157) and were accessed under application number 194410. Analyses using UKB data were performed in the UKB Research Analysis Platform.

### Participant Selection

To minimize population structure bias, the cohort was restricted to participants with self-declared white British ethnicity and similar genetic ancestries based on principal components analysis of the genotypes (UKB field ID 22006). Participants missing >49% of the full UKB-PPP protein abundance data were excluded as outliers based on inspection of the histogram of subject-wise missingness. The remaining participants were included in the downstream analyses.

For each participant, age was taken at the date of enrollment, which was also the date of sample collection (UKB field ID 21022). Sex was acquired from a central registry but in some cases was updated at enrollment by the participant (UKB field ID 31). Pack-years of smoking were calculated based on self-reported history during intake (UKB field ID 20161); lifetime non-smokers, as well as former smokers who started and stopped smoking before age 16, or who started and stopped smoking at the same age and had not smoked for more than six months at enrollment, were assigned zero pack-years.

Diagnosis of AAA was ascertained from linked inpatient health records by presence of ICD-10 codes I71.3 and/or I71.4 (UKB field ID 41270). Both incident and prevalent AAA were included. Similarly, diagnoses of hypertension (HTN) and hyperlipidemia (HLD) were ascertained by presence of ICD-10 codes I10-I1A and E78, respectively. Use of anti-HTN and anti-HLD medications were obtained from assessment center data (UKB field IDs 6177 and 6153), as this was appreciated to be the most complete and current source of prescription data at time of enrollment.

Clinical characteristics of the participants with and without AAA were compared using nonparametric Mann-Whitney U tests for continuous variables (via the wilcox.test function in R) and chi-squared tests for categorical variables (via the chisq.test function in R).

### Protein Abundance Data

The UKB-PPP provides normalized blood plasma protein abundance data for 2,923 proteins per participant. In order to facilitate external validation of the eventual ProRS, analysis was restricted to a candidate list of 275 relevant cardiometabolic proteins based on availability of the proteins in other Olink-based proteomic biorepositories, namely the Swedish Infrastructure for Medical Population-Based Life-Course and Environmental Research (SIMPLER) cohort. Of the 275 candidate proteins, one protein (PCOLCE) was excluded due to outlying missingness in the UKB-PPP participants (>10%). This yielded a final candidate list of 274 proteins for ProRS development (Supplementary Materials S1).

Residual missingness of the protein abundance data was low (approximately 2.2%). The remaining missing values were imputed using a k-nearest neighbors approach with k = 10 via the VIM package in R. To maximize accuracy, all proteins measured in the UKB-PPP were used for distance calculation, except those with missingness >10% (CTSS, NPM1, PCOLCE, TACSTD2, CST1, and GLIPR1; total of 2,917 of 2,923 proteins used). Imputed values were not used for the distance calculation.

### Construction of Proteomic Risk Scores

The proteomic risk score was generated using L1-regularized/Least Absolute Shrinkage and Selection Operator (LASSO) logistic regression implemented with the glmnet package in R. The most well-established clinical risk factors for AAA (age, sex, and pack-years of smoking) were included as unpenalized variables. The remaining variables, including the 274 plasma proteins, diagnosis of HTN and use of anti-HTN medications (and interaction thereof), and diagnosis of HLD and use of anti-HLD medications (and interaction thereof) were penalized. All variables were standardized for modeling; non-standardized regression coefficients are reported in the Supplementary Materials.

A twenty-fold cross validation procedure was used to select two values of the L1-regularization hyperparameter, λ_MIN_ and λ_1SE_, for ProRS construction. Binomial deviance was used as the objective function to minimize. Variables surviving at λ_MIN_, corresponding to the model with the lowest mean binomial deviance in the cross-validation procedure, comprised the “rich” ProRS. A “sparse” ProRS was generated at λ_1SE_, corresponding to the most sparse model with mean binomial deviance within one standard error of the cross-validated estimate at λ_MIN_.

Two additional post-hoc models were created for performance comparison purposes. The first, “Clinical + Smoking,” was composed of the non-protein clinical variables (including smoking) surviving in the “sparse” ProRS obtained at λ_1SE_. The second, “Clinical + MMP12,” was constructed in an identical manner, except the smoking variable was substituted for MMP12 abundance in light of the known association between the two variables.

### Evaluation of Model Performance

Models were assessed for overall performance (via Brier score, a measure of prediction accuracy) and discrimination (via area under the receiver operating characteristic curve; AUROC), in addition to sensitivity, specificity, precision, and recall [17, 18]. This was accomplished using a bootstrapping approach with 2000 replicants. In each iteration, a Brier score was calculated (DescTools package in R), a ROC curve was generated (pROC package in R), the AUROC was recorded, and the sensitivity and specificity were assessed along the ROC curve at the point maximizing Youden’s J statistic. Means and 95% confidence intervals for each of these metrics are reported. Precision and recall curves obtained via the PRROC package in R were bootstrapped in an identical fashion, and mean precision and 95% confidence intervals are plotted for each level of recall.

A Bayesian framework was used evaluate statistical significance and practical equivalence of between-model differences in Brier score and AUROC. This was implemented via the tidyposterior package in R. Both of these metrics were found to be normally distributed by inspection of the quantile-quantile plots. For computational tractability and to ensure sufficient sampling efficiency, a subset of 50 bootstrap samples were used for analysis. Posterior distributions of AUROC and Brier Score were generated for each model using the perf_mod function with 10,000 iterations, including 1000 warmup iterations. These were then used to build posterior distributions for between-model differences via the contrast_models function. Between-model differences were considered statistically significant if greater than 95% of the posterior distribution probability mass was greater than or less than zero. Mean and 95% credible intervals are reported for each pairwise comparison. Practical significance was assessed using a Region of Practical Equivalence (ROPE), defined as a 2% improvement relative to the performance of a null model (i.e. uniform prediction of the negative class). For AUROC, with null performance of 0.5, this was equal to an absolute ROPE of ± 0.01. For Brier score, this was equal to ± 2% of the population-specific AAA prevalence. Posterior distributions with >95% probability mass outside of the ROPE were considered practically equivalent.

Spearman correlation was used to characterize the association between plasma MMP12 abundance and smoke exposure amongst participants with non-zero pack-year smoking history.

### Evaluation of Model Utility

To evaluate practical utility of the ProRS, we computed the Proportion of Cases Followed (PCF), the fraction of AAA cases in the population that would be identified by screening the proportion (q) of the population at highest risk by ProRS [19]. A curve was generated by plotting PCF as a function of q for the included participants. For comparison purposes, the proportion of the population captured by current USPTF/SVS screening recommendations (i.e. ever-smokers aged ≥65 at enrollment) was also calculated [1, 2]. The USPTF/SVS screening guidelines were specifically operationalized as ever-smokers aged ≥65 at enrollment; family history of AAA was not included in the definition as it was not captured in UKB. The number needed to screen (NNS) to detect one AAA was computed for both the proportion of the population captured by the operationalized USPTF/SVS criteria as well as an equal proportion of the population at highest risk by ProRS by dividing the number of individuals screened by the number of captured aneurysms.

### Relationship between MMP12 and Time to Aortic Rupture or Repair

To explore the relationship between plasma MMP12 abundance and time to aortic rupture or aortic aneurysm repair amongst participants with incident AAA (i.e., AAA diagnosed after enrollment), we constructed a Cox proportional hazards model with age and sex as covariates. This was accomplished using the survival package in R. Time to event was defined as the number of days between sample collection and earliest occurrence of either aortic rupture (as defined by ICD-10 code I71.3) or aortic aneurysm repair (as defined by OPCS4 codes, see Supplementary Materials S2). The remaining participants were right-censored at 31 March 2023, the date up to which UKB estimates that ICD/OPCS data is reasonably complete, or date of death, whichever occurred first.

### External Assessment of MMP12 Predictive Utility

To further characterize the utility of plasma MMP12 abundance for prediction of AAA, we assessed the performance of an age + sex + smoking + MMP12 model over an age + sex + smoking null model in the SIMPLER cohort. Use of the SIMPLER data was approved by the Swedish Ethical Review Authority, and detailed information on the cohort and quality controls have been described previously. Similar to UKB-PPP, plasma protein abundance data were obtained using an Olink assay. For each participant, age and sex were obtained from questionnaire data. Smoking history was measured as binary “ever” or “never,” as opposed to the continuous pack-years variable used in the UKB analyses. Presence of AAA was ascertained from the nationwide Swedish Patient Register based on presence of ICD-10 primary diagnosis codes I71.3 or I71.4. Plasma MMP12 abundance levels were Z-score normalized prior to analysis, and all models were constructed using the glm function in R. Performance of the two SIMPLER models was assessed and compared in an identical manner to the UKB analyses. PCF and NNS for the MMP12-containing model and current USPTF/SVS screening recommendations were computed in the SIMPLER population as previously described.

## Results

### Study Population

A total of 52,998 UK Biobank Pharma Proteomics Project (UKB-PPP) participants had plasma protein profiling performed at time of enrollment. After exclusion of 9,251 individuals with non-white British ethnicity and/or dissimilar genetic ancestries, as well as 6,768 individuals with outlying subject-wise protein data missingness, 36,979 individuals (70%) were retained for downstream analyses. Of the included participants, 267 (0.72%) had AAA. The majority of AAA was incident (242 of 267; 90.6%), and the mean time of AAA diagnosis was 7.4 ± 4.9 years after enrollment. Participants with AAA were older (63.9 versus 57.2 years; p < 0.001), more likely to be male (84.3 versus 45.8 percent; p < 0.001), and had higher pack-years of smoking (23.4 versus 7.4 pack-years; p < 0.001). Participants with AAA were also more likely to have HTN (79.4 versus 33.5 percent; p < 0.001), take medication for HTN (52.4 versus 21.8 percent; p < 0.001), have HLD (53.9 versus 16.9 percent; p < 0.001), and take medication for HLD (57.7 versus 18.4 percent; p < 0.001). A tabular description of the cohort is included in the Supplementary Materials (S3). After imputation, protein abundance data were available for each of the 274 candidate proteins for each subject.

### Development of Proteomic Risk Scores

Two candidate ProRS were generated using a variable selection procedure. Age, sex, and pack-years of smoking were included by default in both models. The first ProRS (termed “LASSO Rich”) was constructed to maximize classification performance. It contained 71 proteins, the three default variables, and terms for HTN, HLD, and medication for HLD. The second ProRS (termed “LASSO Sparse”) was constructed to balance model sparsity and model performance, and contained only one plasma protein – matrix metalloproteinase 12 (MMP12). The remaining terms in this model included the three default variables, terms for HTN and HLD, and an interaction term between diagnosis of HLD and medication for HLD. Within the LASSO Sparse model, male sex and MMP12 abundance had the largest regularized regression coefficients. The complete variable selection procedure is summarized in Figure 1, and a full listing of surviving variables and regularized coefficients for the LASSO models are included in the Supplementary Materials (S4 and S5).

**Figure 1:**
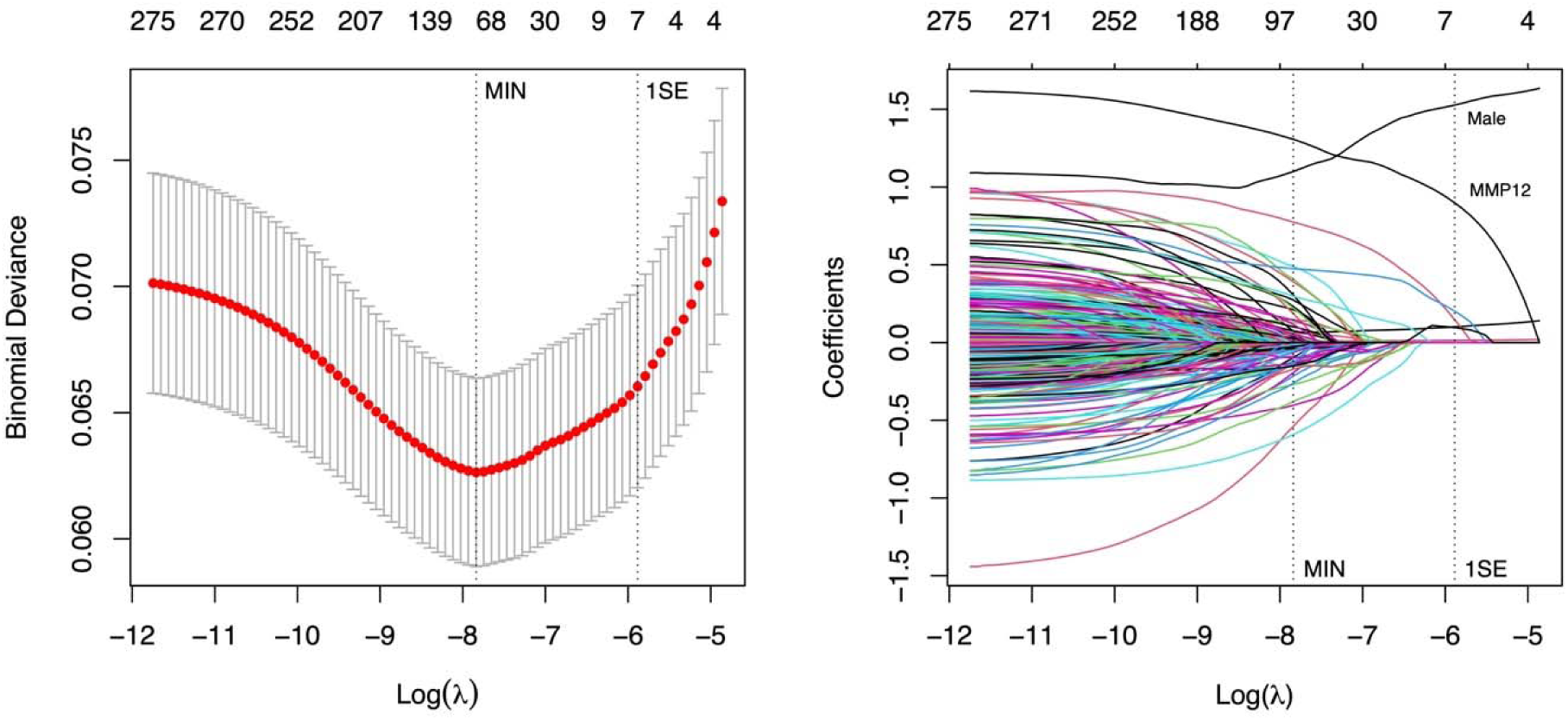
LASSO Regularization Procedure. LEFT: LASSO ProRS models were constructed at the value of λ minimizing the binomial deviance (λ_MIN_; LASSO Rich) and the value of λ with mean binomial deviance within one standard error of the estimate at λ_MIN_ (λ_1SE_; LASSO Sparse). RIGHT: Surviving variables at each value of λ. Male sex and MMP12 abundance had the highest regression coefficients in the LASSO Sparse model at λ_1SE_.

Two comparison models were generated based on the ProRS results: “Clinical + Smoking” included all variables in LASSO Sparse except for MMP12; “Clinical + MMP12” was identical to “Clinical + Smoking,” except that the smoking variable was substituted for MMP12 abundance, in view of known associations between MMP12 and risk of AAA among smokers [20].

### Evaluation of Model Performance

Bootstrapped estimates of area under the receiver operating characteristic curve (AUROC), sensitivity, specificity, and Brier scores for each of the two ProRS models and two comparison models are shown in Figure 2 and reported in the Supplementary Materials (S6). All between-model differences in AUROC and Brier score were statistically significant in the Bayesian analysis. Furthermore, except for the LASSO Sparse to Clinical + MMP12 contrast, none of the between-model differences in AUROC or Brier were practically equivalent (region of practical equivalence [ROPE] ± 0.01 for AUROC and ± 1.4 × 10^-4^ for Brier score). Full results of the between-model analysis are included in the Supplementary Materials (S7).

**Figure 2:**
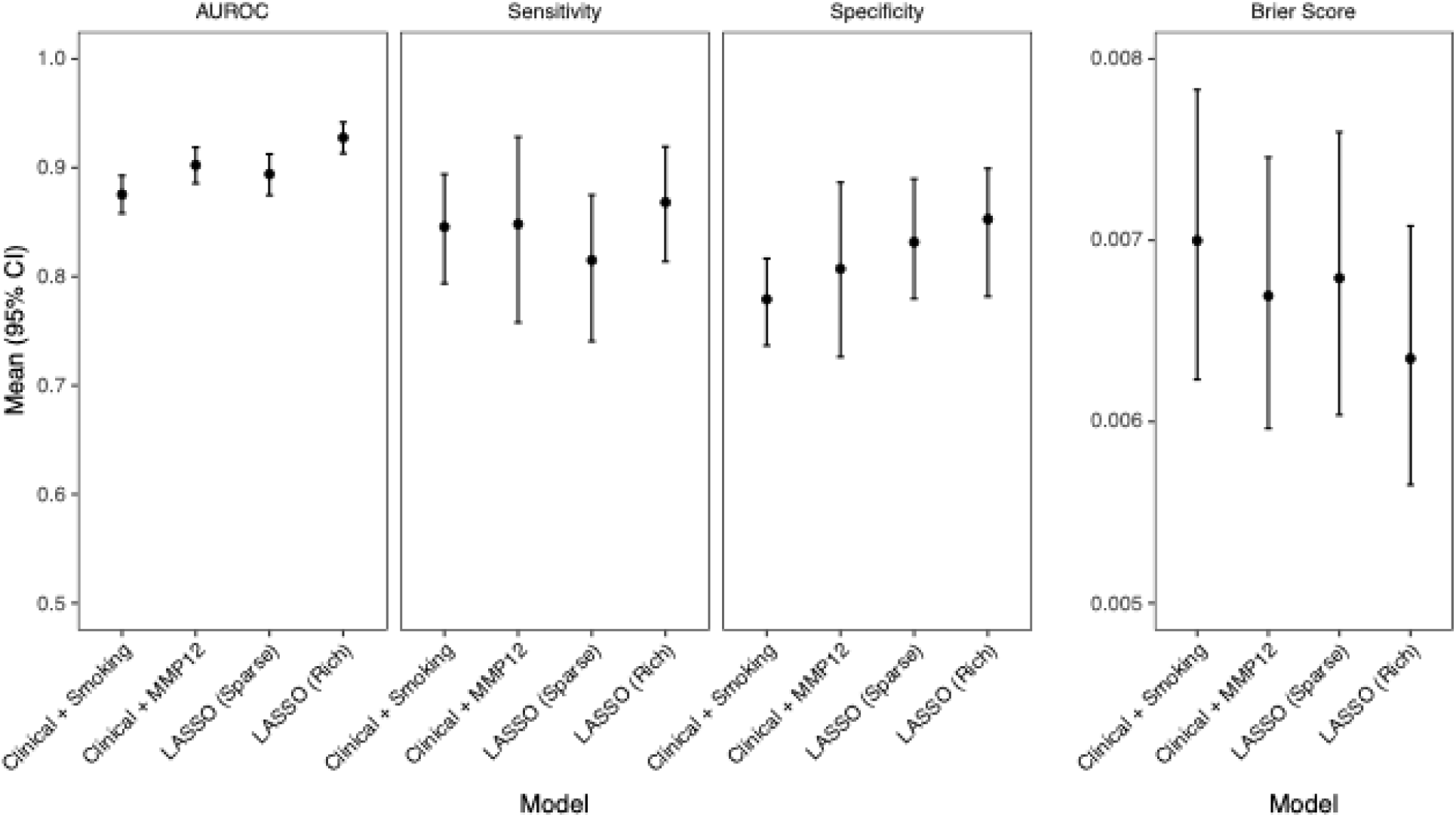
Model Performance. Estimates of area under the receiver operating characteristic curve (AUROC; LEFT), sensitivity (MIDDLE LEFT), specificity (MIDDLE RIGHT), and Brier score (RIGHT) along with 95% confidence intervals obtained using 2000 bootstraps.

The LASSO Rich model with 71 plasma proteins had the highest overall performance (mean Brier score 6.3 × 10^-3^, 95% CI 5.7 × 10^-3^ - 7.1 × 10^-3^) and best discrimination (mean AUROC 0.928, 95% CI 0.913 - 0.942). Of particular interest, the LASSO Sparse model had higher overall performance and better discrimination than Clinical + Smoking, an identical model without MMP12 (difference in Brier score 2.1 × 10^-4^, 95% CrI 2.0 × 10^-4^ - 2.3 × 10^-4^; difference in AUROC 0.021, 95% CrI 0.020 - 0.022). Furthermore, using MMP12 as a surrogate for pack-years (Clinical + MMP12 model versus Clinical + Smoking model) also improved overall performance and discrimination (difference in Brier score 3.2 × 10^-4^, 95% CrI 3.1 × 10^-4^ - 3.4 × 10^-4^; difference in AUROC 0.029, 95% CrI 0.027 - 0.030). Interestingly there was only a relatively low correlation between plasma MMP12 level and pack-years amongst smokers (Spearman ρ = 0.323; p < 0.001). Estimates of sensitivity and specificity were generally high for all models with most means in the range of 0.8 – 0.9, although with comparatively large confidence intervals relative to the estimates of AUROC.

Bootstrapped precision-recall curves for the LASSO Sparse, LASSO Rich, and Clinical + Smoking models are shown in Figure 3. The LASSO Sparse model with MMP12 had higher precision than the Clinical + Smoking model (without MMP12) at moderate levels of recall.

**Figure 3:**
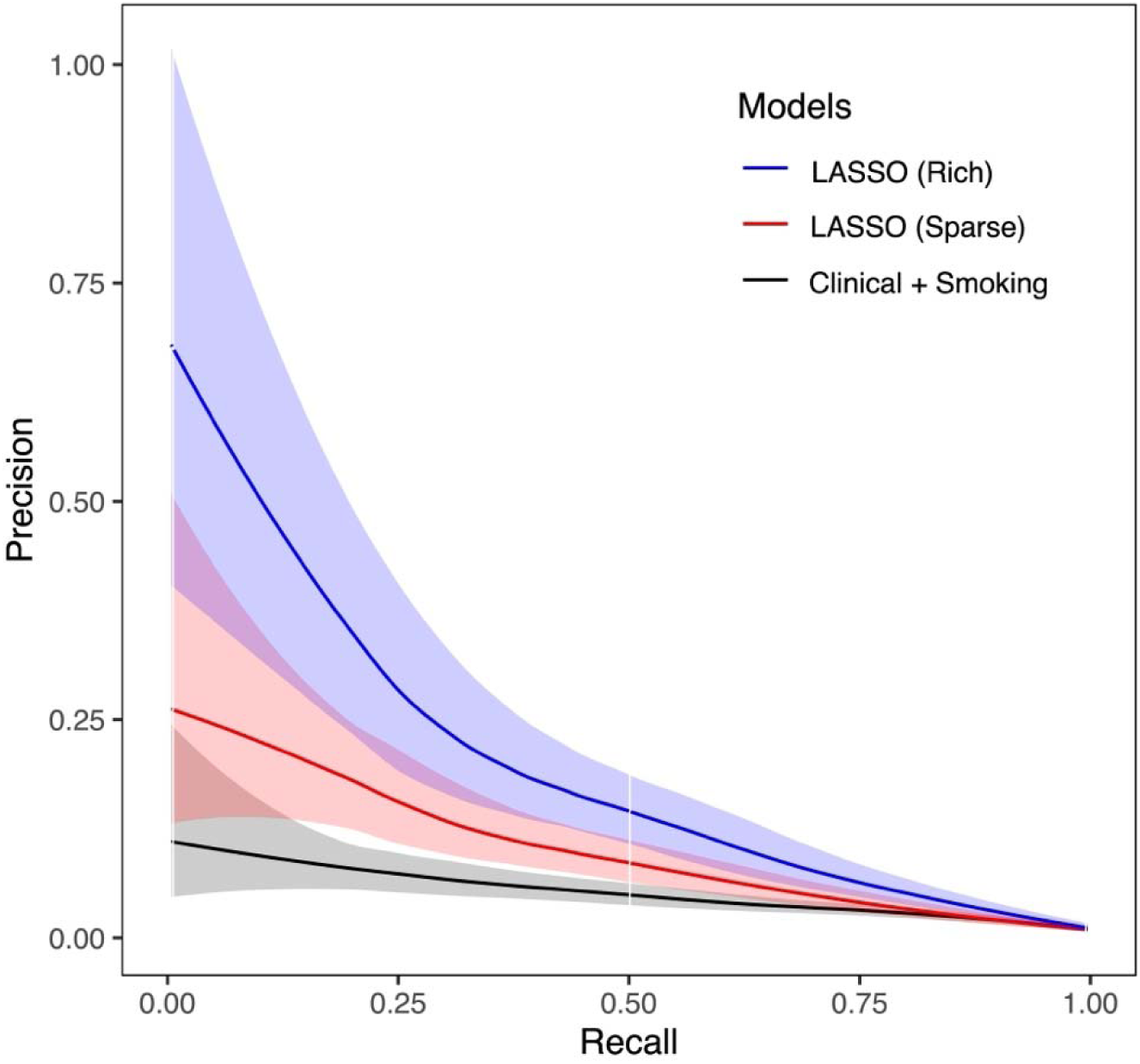
Precision-Recall Curves. The shaded areas represent 95% confidence intervals for precision at a given level of recall obtained via 2000 bootstraps of the precision-recall curve.

### Evaluation of Model Utility

The performance of the LASSO Sparse ProRS as a risk stratification tool was compared to the operationalized USPTF/SVS AAA screening recommendations by computing the respective proportion of cases followed (PCF; see Methods). A plot of PCF as a function of the proportion of the population screened at highest risk by the ProRS is presented in Figure 4. Within the UKB-PPP cohort, traditional USPTF/SVS screening recommendations captured 4.6% of the population and 30% of AAA cases, whereas screening 4.6% of the population at highest risk by ProRS captured 52% of cases. To capture 30% of AAA cases using ProRS, only 1.6% of the population must be screened. Within the 4.6% of the population captured by USPTF/SVS recommendations or at highest risk by ProRS, the number needed to screen to detect one aneurysm was substantially lower using the ProRS (21 versus 12 individuals).

**Figure 4:**
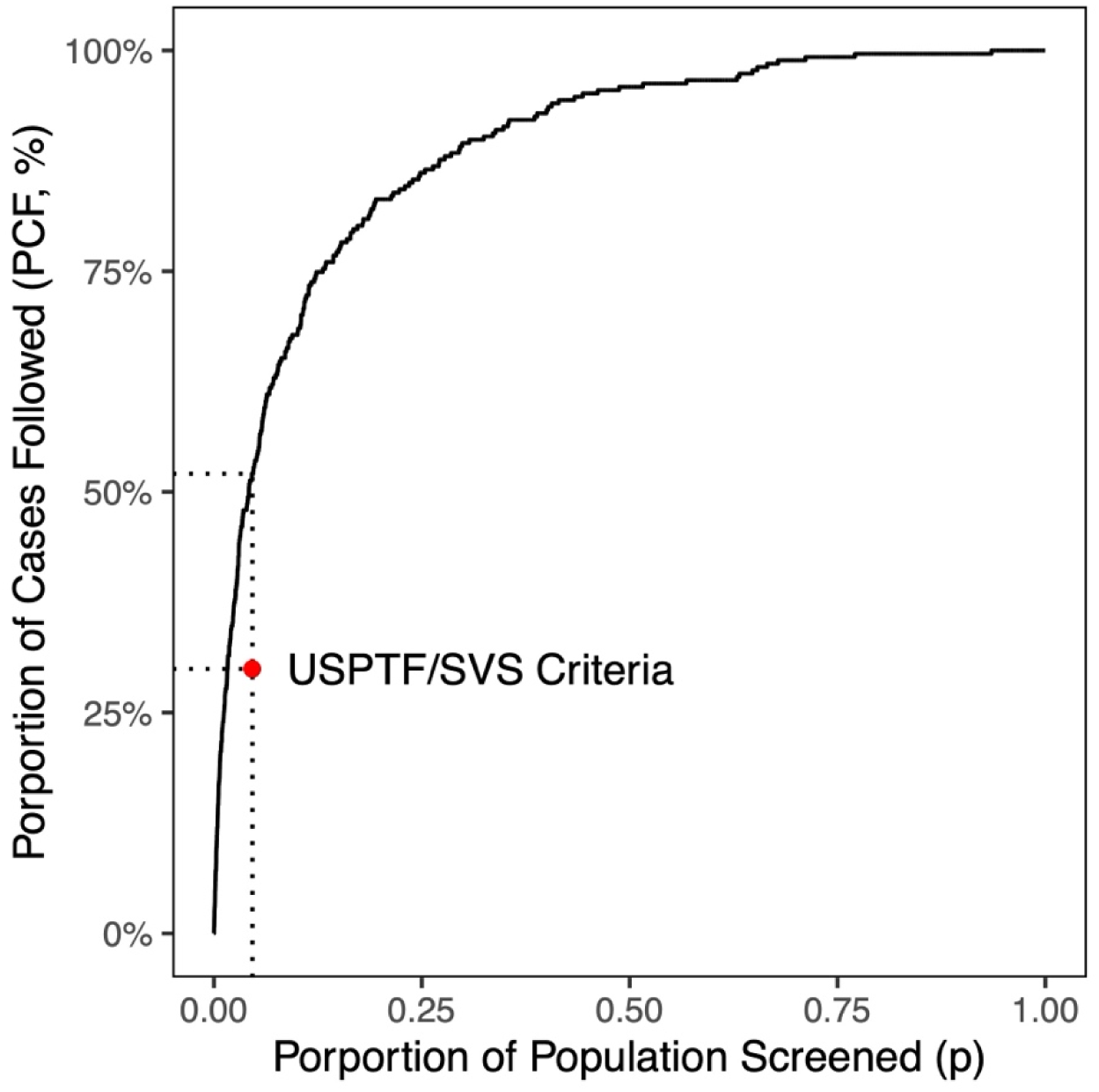
Proportion of Cases Followed (PCF) as a function of the proportion of the population screened at highest risk by the LASSO Sparse ProRS in the UKB-PPP cohort. The red dot represents the performance of the USPTF/SVS screening criteria in the UKB-PPP population (4.6% of population screened, capturing 30% of AAA).

### Relationship between MMP12 and Time to Aortic Rupture or Repair

To establish the relationship between plasma MMP12 abundance and time to aortic rupture or repair amongst participants with incident AAA (N = 242), we constructed a Cox proportional hazards model with age and sex as covariates. A total of 79 individuals (33%) experienced rupture or underwent repair. Within this population, plasma MMP12 abundance was independently associated with time to rupture or repair (hazard ratio 1.86, 95% confidence interval 1.39 – 2.50, p < 0.001). Full regression results are included in the Supplementary Materials (S8).

### External Assessment of MMP12 Predictive Utility

To further characterize the predictive utility of plasma MMP12 abundance for prediction of AAA, performance of a model containing age, sex, smoking, and MMP12 was compared to a null model (age, sex, and smoking alone) in an external population of 12,314 individuals from SIMPLER. Compared to UKB-PPP, the SIMPLER cohort had higher rate of AAA (2.20% versus 0.72%), older participants (mean ± standard deviation; 71.6 ± 6.8 versus 57.3 ± 8.1), and higher proportion of males (61.5% versus 46.1%). Proportion of ever-smokers was similar between the two populations (35.3% versus 31.1%). Within the SIMPLER cohort, the model containing MMP12 had higher overall performance compared to the null model without MMP12, a difference that was statistically significant (difference in Brier score 1.1 × 10^-3^, 95% CrI 1.0 × 10^-3^ – 1.2 × 10^-3^) but practically equivalent (ROPE ± 4.4 × 10^-2^; Supplementary Materials S9). The MMP12-containing model also had higher discrimination, which was both statistically significant (difference in AUROC 0.084, 95% CrI 0.081 - 0.087) and practically non-equivalent (ROPE ± 0.01). USPTF/SVS screening recommendations captured 20% of the population and 54% of AAA, corresponding to an NNS of 17 individuals, whereas screening the same proportion of the population at highest risk by the MMP12-containing model captured 69% of AAA with a reduced NNS of 13 individuals.

## Discussion

In this large cross-sectional analysis of nearly 37,000 UKB-PPP participants, we developed a sparse ProRS in which a single measurable plasma protein (matrix metalloproteinase 12) plus well-established clinical risk factors substantially improved the prediction of AAA over clinical factors alone. Plasma MMP12 also increased discrimination of AAA in an external cohort, and was independently associated with time to rupture or repair for incident AAA.

MMP12 has well-established elastolytic functions in the extracellular matrix and is known to be implicated in AAA pathogenesis [21]. Previous studies have demonstrated upregulation of MMP12 in both murine models of AAA [22, 23] as well as in human AAA [24, 25, 26]. The emergence of MMP12 as an important proteomic predictor of AAA is therefore biologically plausible and likely to be reflective of the underlying pathogenesis. These results help inform the potential biological relationship between smoking, MMP12, and AAA, and have implications for future screening strategies to detect AAA.

The relationship between smoking and MMP12 abundance warrants additional consideration. Smoking-induced vascular inflammation and oxidative stress is thought to have activating effects on cellular sources of matrix metalloproteinases, such as macrophages and endothelial cells, as well as additional activating effects on gene transcription, zymogen activation, and impairment of MMP inhibition [27]. Smoking-induced increases in tissue MMP12 expression are also known to be associated with atherosclerotic disease [28]. Furthermore, a Mendelian randomization study has identified MMP12 as an important mediator of the association between smoking and AAA [20]. In our analysis, we found that using MMP12 instead of self-reported pack-years of smoking improved prediction of AAA in a clinical-variable-only model, with only a modest correlation between MMP12 level and pack-years amongst smokers. This implies that MMP12 has higher discriminatory utility than pack-years, and by extension, considering the strong biological and epidemiologic association between smoking and AAA, may be a more accurate reflection of cumulative smoking history than a patient’s self-reported exposure.

From a screening perspective, compared to current USPTF/SVS recommendations, screening patients at highest risk by sparse ProRS nearly doubled the proportion of AAA cases followed in the UKB-PPP cohort. Although the rich ProRS incorporating 71 proteins had the best performance, the proposed sparse ProRS has the key advantage of relatively practical clinical implementation, given that it only requires measurement of a single plasma protein. It could plausibly be employed for targeted screening or individualized risk stratification at the primary care level. Our results suggest the sparse ProRS could have a role in early detection of AAA, noting strong predictive performance in a population that, on average, had plasma protein measurement performed more than seven years prior to establishment of a clinical diagnosis. Furthermore, we identified associations between plasma MMP12 level and time to aortic rupture or repair that may carry potential prognostic utility. Implementation of the ProRS to fine-tune screening efforts based on individual biological risk has the potential to improve healthcare resource allocation and reduce health system inefficiencies associated with one-size-fits-all guidelines.

The present study has several strengths, including a large sample size, direct measurement of plasma protein abundance, robust cross-validation and bootstrapping procedures, and external validation of MMP12 predictive utility. There are several key limitations. First, in order to increase sample size, minimize cohort imbalance, and reduce risk of model overfitting, no differentiation was made between incident and prevalent AAA for model training. Future studies with a greater number of positive cases would be required to generate a ProRS specifically designed to predict incident AAA or risk of AAA rupture. Second, it should be noted that this analysis was restricted to participants with white British ethnicity, potentially limiting generalizability. Additional analyses are needed to characterize the plasma proteomic profile of AAA in more diverse populations. Third, the cross-sectional nature of this study is fundamentally subject to ascertainment biases, particularly survivorship biases, which may skew the proteomic signature of disease towards more slowly-progressive or late-developing cases.

Lastly, given the lack of longitudinal data, it is unknown how ProRS or MMP12 level might change over time, or change in relation to time of onset of AAA. Ultimately, additional studies will be needed to further characterize the performance and interpretation of the ProRS and compare biologically-informed screening strategies to conventional approaches.

### Conclusions

A parsimonious ProRS incorporating a single matrix metalloproteinase known to be implicated in AAA disease pathogenesis improved prediction of AAA over clinical factors alone in a broad clinical population.

## Data Availability

All data produced in the present study are available upon reasonable request to the authors.

https://www.ukbiobank.ac.uk

https://www.simpler4health.se

## Acknowledgements

The UKB analyses were conducted using the UK Biobank Resource under application number 194410. We acknowledge SIMPLER for data access, and we would like to thank Anna-Karin Kolseth and Niclas Håkansson for assistance. This publication does not represent the views of the Department of Veterans Affairs or the United States Government.

